# Spiral Septal Morphology Distinguishes Arrhythmic from Idiopathic DCM and Links to Prognosis

**DOI:** 10.64898/2026.02.17.26346514

**Authors:** Clint Asher, Crispin Musicha, Gabriel Balaban, Reza Razavi, Gerry Carr-White, Pablo Lamata

## Abstract

**BACKGROUND:** Dilated cardiomyopathy (DCM) presents a highly heterogeneous spectrum, including a familial subset with elevated arrhythmic risk. Traditional demographic and imaging markers, such as late gadolinium enhancement, have been inadequate for identifying high-risk patients before arrhythmic events. Remodelling of the interventricular septum—central to ventricular mechanics and conduction—may offer improved risk stratification.

**OBJECTIVES:** To identify differences in left ventricular (LV) morphology between arrhythmic and idiopathic dilated cardiomyopathy (aDCM vs iDCM), and to identify LV remodeling patterns that link to adverse outcomes.

**METHODS:** Three-dimensional LV shape models were constructed from end diastolic cardiovascular magnetic resonance images of 102 individuals subdivided by their idiopathic or arrhythmic subgroup allocation. A statistical shape model was built using principal component analysis. A linear discriminant analysis determined shape features of the arrhythmic subgroup and increased composite arrhythmic outcome of sudden cardiac death, aborted sudden cardiac death, and sustained ventricular tachycardia.

**RESULTS:** The idiopathic DCM group displayed larger mass, length, diameter, mass to volume ratio, and a mild spiral pattern of thicker septal walls (p=0.004). The arrhythmic DCM group displayed a more conical (wider basal and mid wall to apical diameter) LV, and the lack of the spiral septal morphology was the most significant feature (p=0.006) to identify subjects that had the composite arrhythmic outcome.

**CONCLUSION:** The LV morphology derived suggests a differentiation of arrhythmic DCM patients beyond size, function and LGE presence. This was distinctive and captured shape features that suggest alternate mechanisms for arrhythmic risk linked to a pattern of remodeling.

**Graphical Abstract:** Assessing LV morphology signature of arrhythmic DCM phenotype

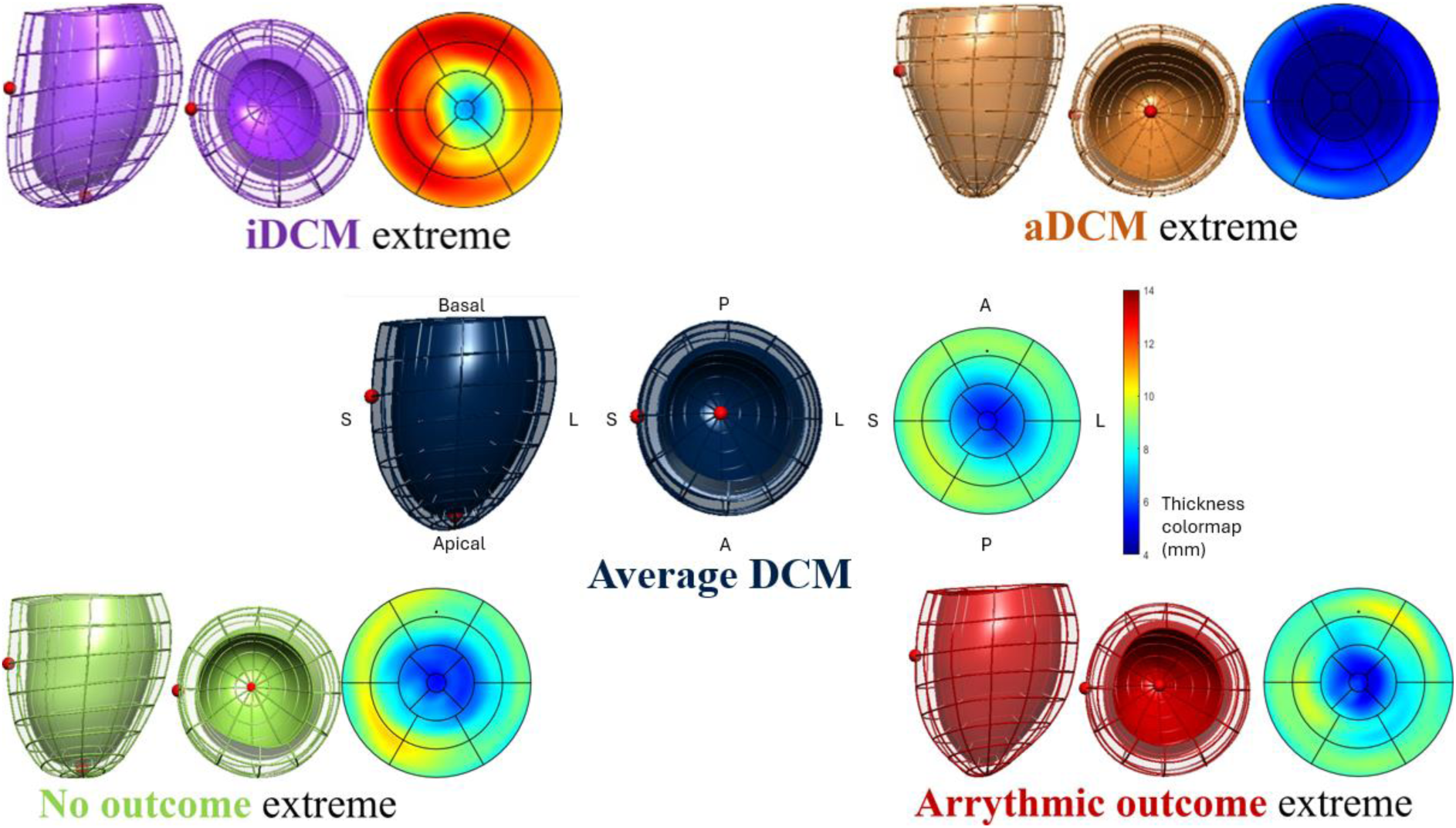

## Introduction

Ventricular arrhythmias are a major clinical concern in non-ischemic dilated cardiomyopathy (DCM) (1). In recent years, a familial subset with an arrhythmic phenotype has been recognized (2). This is one of the causes of arrhythmogenic cardiomyopathy (ACM), whereby ventricular arrhythmias may occur despite lack of significant left ventricular (LV) eccentric remodeling (2,3). As such, this arrhythmic DCM (aDCM) subset can have a clinical course that appears to be more malignant, raising the risk of adverse outcomes, therefore warranting closer monitoring and potentially earlier intervention (2,4).

Currently, the identification of aDCM relies on comprehensive assessment for higher-risk clinical features and appropriate genetic testing (2). However, substantial genetic heterogeneity in DCM and limitations in early genotype-phenotype correlations mean accurate clinical characterization is often problematic (5–9). Given the expectation that at least a quarter of presumed idiopathic DCM (iDCM) will also have an underlying genetic predisposition (6,10), some of these patients may have similar high-risk features and share common pathways in their risk of arrhythmic events (11). This highlights the increasing urgency for novel diagnostic pathways to better identify and characterize these patients.

Recent work suggests shape analysis, which captures geometric features beyond simple volume and mass metrics, may provide sensitive indicators for arrhythmia prediction in fibrotic DCM patients (12), however its role in those with arrhythmic risk despite mild LV eccentric remodeling and/or without fibrosis is unclear and a present need.

In this study, we sought to analyze aDCM compared to iDCM using enhanced three-dimensional (3D) descriptors of shape and to identify LV remodeling patterns that link to adverse outcomes.

## Methods

The data underlying this article are available in open access in the FigShare repository (DOI: tobecreated), and tools are available upon request in GitHub.

### Study Population & Consent

Patients were enrolled consecutively following clinical evaluation in the heart failure and inherited cardiac services at Guys’ and St Thomas NHS Trust, with consent to clinical cardiovascular magnetic resonance imaging (CMR) scanning for further analysis. Patients were enrolled between 2015 and 2020 and gave written informed consent to follow up on outcomes. Patients had non-ischemic DCM, characterized by varying degrees of reduced LV ejection fraction (LVEF) compared with published reference ranges normalized for age and sex (13). Included are those with impaired function i.e. LVEF≤50% in the absence of increased LV end-diastolic volume to capture the early disease phenotype. This is formally recognized as hypokinetic non-dilated cardiomyopathy and adds further to the potential benefit offered by early shape changes (14). Patients were excluded if they had significant valvular disease, hypertensive heart disease or coronary artery disease that would be commensurate with the degree of LV impairment. All secondary causes of DCM were excluded.

The aDCM subset was defined clinically as those with an increased risk of arrhythmic events as set out in the task force documentation for ACM (2,15). We considered those with familial DCM (i.e., at least another first degree relative with the DCM phenotype), and subsequently to meet the aDCM inclusion also had one or more of: preceding history of unexplained syncope; family history of sudden cardiac death less than forty years of age; preceding history of non-sustained ventricular tachycardia (NSVT), that was characteristically monomorphic; more than one thousand ventricular ectopics every twenty-four hours on

Holter recording; non-ischemic late gadolinium enhancement (LGE) on CMR in two orthogonal planes. None of these subgroups had a personal history of cardiac arrest prior to enrolment.

### Clinical Follow-up and Composite Arrhythmic end-point

Patients had clinical follow up from the point of their CMR using questionnaires, telephone interviews and review of health records. An arrhythmic outcome following CMR was documented as a composite of: aborted sudden cardiac death, sustained VT, or sudden cardiac death.

### Cardiac magnetic resonance acquisition and analysis

The CMR studies consisted of consecutive biventricular short axis (SAX) slices acquired at rest during breath-hold, ECG-gated cine SSFP acquisition, performed at 1.5 or 3 Tesla.

End diastolic LV contours and RV endocardium were semi-automatically segmented (CVi42, Circle Cardiovascular imaging Inc.) with expert manual correction, as clinically required. As per clinical practice, the papillary muscles and trabeculae were included within the blood pool contours. Areas of LGE were identified with semi-automated full-width at half maximum approach by an independent CMR expert blinded to patient outcomes.

### Statistical shape model of the left ventricle

Personalized 3D anatomical models of the LV at end diastole were automatically generated by fitting a template mesh into their corresponding LV segmented anatomy as illustrated in Figure 1 using methods previously described (16,17). Fitting accuracy was assessed by a measure of the distance from mesh surface to contours.

**Figure 1:**
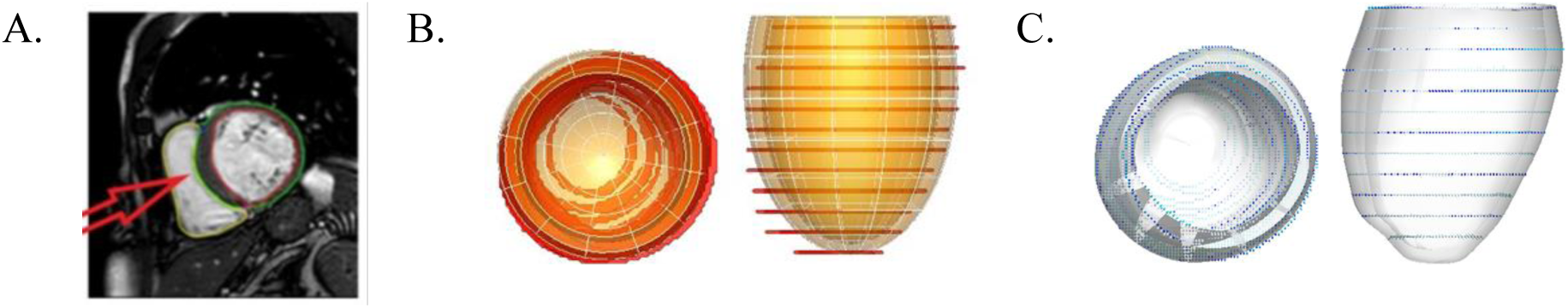
Overview of the automated mesh customization. **Panel A**: CMR imaging with biventricular segmentation, with red arrow pointing to the center of mass of the right ventricle (RV). **Panel B**: Two complementary views of the idealized ellipsoid (transparent orange) template mesh overlaid to the left ventricular contours (red) available from the SAX acquisition planes. **Panel C**: Two complementary views of the personalized computational mesh (semi-transparent white) overlaid to the SAX segmentation contours color-coded by fitting error measured in mm, from 0mm(blue) to 1mm or more(red).

The LV meshes were spatially aligned by their center of mass and by orientation defined by two directions, the perpendicular to the SAX planes and the line joining the center of mass of the LV and RV blood pools. A statistical shape model of the cohort, describing the mean shape and its 3D modes of variation was built with Principal Component Analysis (PCA). As a result, each of the meshes was described as a set of coefficients, one per PCA mode.

The 3D shapes were then investigated to assess differentiation of (1) the two clinical subgroups (iDCM vs. aDCM) and (2) patients who met the composite outcome vs no-outcome. A linear discriminant analysis (LDA) was utilized to combine PCA modes as described previously (18). Modes were selected using least absolute shrinkage and selection operator (LASSO feature selection algorithm).

### Statistical Analysis

Normality was tested using the Shapiro-Wilk test. Baseline clinical data and CMR variables are presented as mean± standard deviation by group (iDCM vs aDCM) if normally distributed and compared with an unpaired two tailed Student’s t test. Otherwise, data is presented as median (interquartile range) and compared using the Mann-Whitney U test. Categorical and binary variables were summarized by frequency (percentage) by group. Chi-squared tests were used to assess if there were significant differences in demographic characteristics between the two groups.

For determining differences between shape coefficients for each group along each axis (both the PCA and LDA), a two tailed Student’s t-test was used. A p-value <0.05 was considered statistically significant for all comparisons. Discriminatory performance of shape features was assessed using the area under the receiver operating characteristic (AUC). To minimize overfitting, a cross-validation test of performance using a leave one out strategy was utilized.

All analyses were performed in IBM SPSS Statistics Software Version 28.0.1.1 (14) (International Business Machines, Armonk, New York, USA) and MATLAB (MathWorks, Natick, MA, USA, R2022A).

## Results

### Study population characteristics

102 patients were recruited with baseline characteristics summarized in Table 1. This resulted in a comparison of 50 clinically derived aDCM patients and 52 idiopathic DCM (iDCM) patients, the latter who had no other features meeting local consensus criteria for genetic testing. The iDCM group had a higher proportion of men (79%) versus the aDCM group (48%), p = 0.001.

**Table 1.**
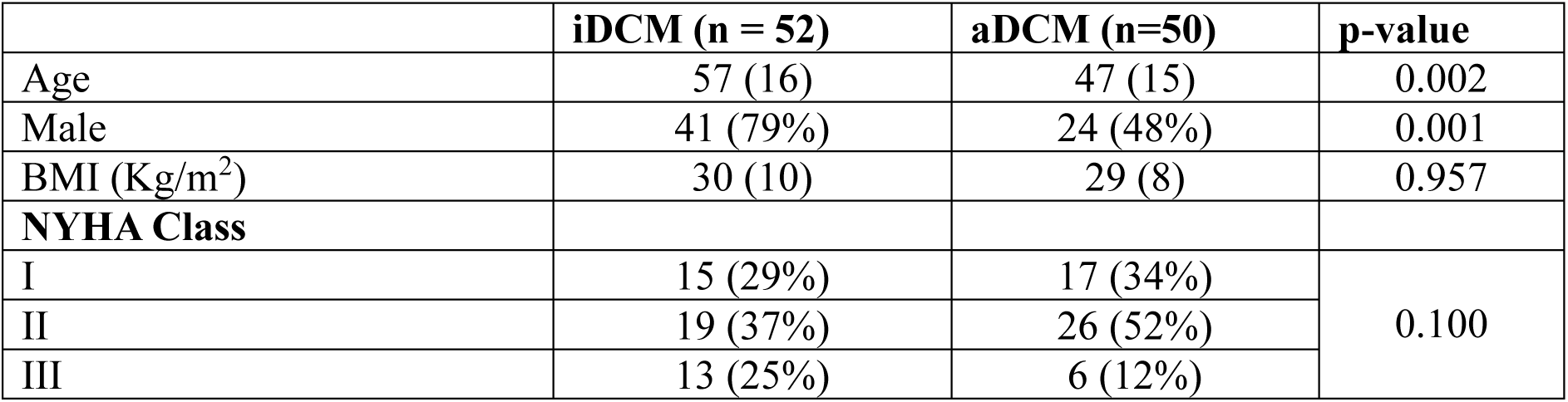
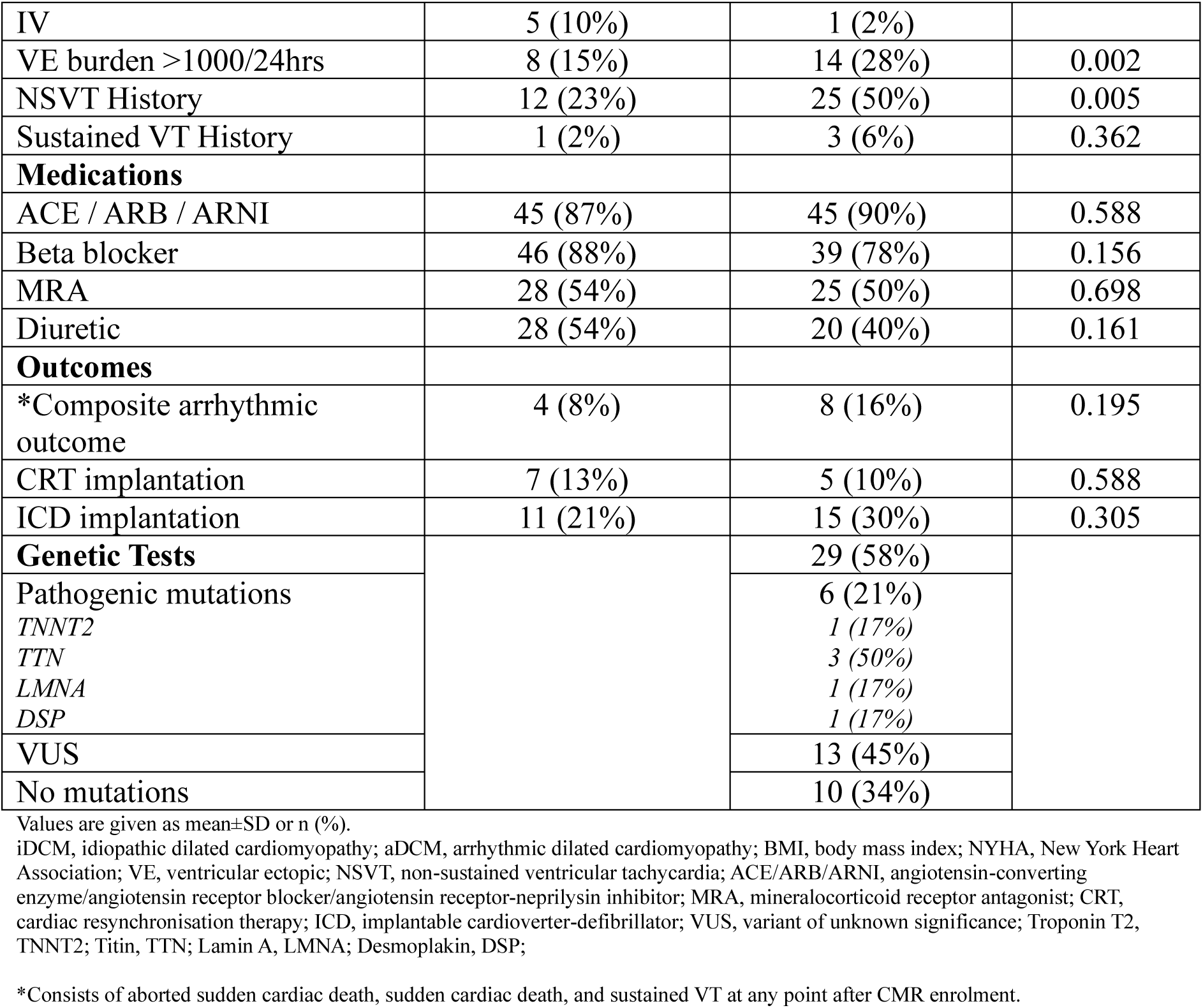
Study population baseline characteristics.

The aDCM subgroup had a significantly higher burden of ventricular ectopics (28% compared with 15% in iDCM subgroup) and non-sustained VT (50% compared with 23% in iDCM subgroup) prior to CMR enrolment (p<0.05 for both). Genetic testing was conducted in 29 of the suitable aDCM patients revealing six pathogenic mutations and thirteen variants of unknown significance (VUS).

The number of aDCM patients meeting the composite arrhythmic end point was 16% compared to 8% encountered in the iDCM subgroup, see Table 1.

### CMR characteristics

The CMR characteristics are summarized in Table 2. aDCM participants had a higher LVEF when compared to their iDCM counterparts, (LVEF 40% vs 34.0%, p = 0.03). Presence of non-ischemic LGE was equally prevalent (42% vs 37%, in the aDCM and the iDCM group respectively). The regional distribution of LGE is reported in Supplementary Section 1. LV wall motion abnormality (WMA) was more prevalent in the aDCM group (22% vs. 0%, p < 0.001).

**Table 2.** Study population CMR characteristics.

|  | iDCM (n = 52) | aDCM (n=50) | p-value |
| --- | --- | --- | --- |
| LVEDV (ml) | 240 (196-295) | 217 (184-274) | 0.214 |
| LVESV (ml) | 155 (121-226) | 129 (104-184) | 0.072 |
| LVEF (%) | 34 (24-39) | 40 (26-46) | 0.03 |
| RVEDV (ml) | 183 (139-214) | 173 (146-208) | 0.459 |
| RVESV (ml) | 92 (73-121) | 89 (65-116) | 0.451 |
| RVEF (%) | 46 (38-54) | 49 (40-55) | 0.209 |
| **WMA** |  |  |  |
| LV WMA | 0 | 11 (22%) | <0.001 |
| RV WMA | 0 | 2 (4%) | 0.238 |
| **LGE** |  |  |  |
| Non-isch | 19 (37%) | 21 (42%) | 0.161 |

Values are given as median (25–75% percentiles) or n (%).
iDCM, idiopathic DCM; aDCM, arrhythmic DCM; LV, left ventricle; RV, right ventricle; EDV, end diastolic volume; ESV, end systolic volume; EF, ejection fraction; WMA, wall motion abnormality; LGE, late gadolinium enhancement; Non-isch, non-ischaemic.

### A statistical shape model of the LV with DCM

All 102 meshes were successfully reconstructed from SAX image contours, achieving a good fitting accuracy with the average error being smaller than the voxel size (error = 1.07 +/- 0.37 mm). The statistical shape model concentrated 91.7% of the variance in shape in the first 10 modes. An illustration, quantification of variance, and interpretation of the 25 first PCA modes are provided in the Supplementary Section 2.

### Shape differences between aDCM and iDCM

Two PCA modes captured differences between aDCM and iDCM groups, modes 3 (p= 0.013) and 24 (p = 0.005), see Figure 2. Mode 3 captured a left to right axis shift in the 3D orientation with an increase in thickness, and Mode 24 captured a spiral pattern of septal wall thickening, both associated with the iDCM group.

**Figure 2:**
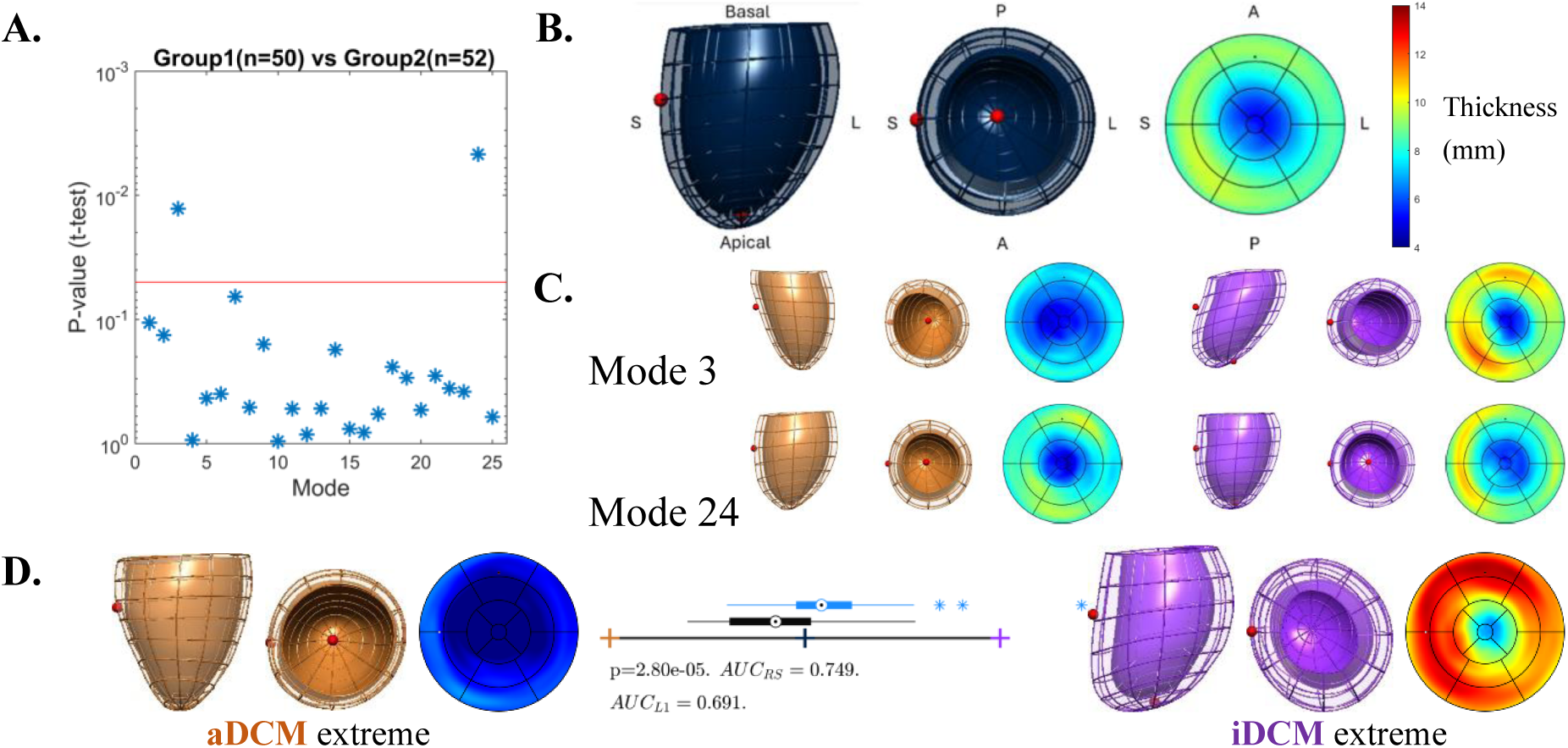
Shape differences captured between the iDCM (n=52) and aDCM (n=50) groups. **Panel A:** Result of the T-test comparing the two groups, where modes 3 and 24 are above the threshold of significance set at p=0.05. **Panel B:** Illustration of the average LV across the 102 DCM cases of this study, with two 3D views and bullseye plot of wall thickness (jet colourmap, in mm) – A: Anterior; P: Posterior; S: Septal; L: Lateral. **Panel C:** Illustration of the two modes (3 and 24) by their extreme shapes (orange as the aDCM extreme and velvet as the iDCM extreme). **Panel D:** Illustration of the changes encoded by the LDA axis on the first 10 PCA modes, with corresponding boxplots of the distribution of aDCM (black) and iDCM (blue) groups along this LDA axis.

Beyond these salient features, the LDA over the first 10 PCA modes showed that the LV of the iDCM subgroup displayed relatively thicker walls and a more globular shape (as opposed to a conical shape of aDCM), see Panel D in Figure 2. Modes 1,2,3,7 and 9 were selected by the LDA using the LASSO gradual aggregation and achieved an AUC of 0.691 in cross-validation.

Mesh-derived geometrical metrics revealed that the aDCM LV had smaller mass, shorter length and diameter, smaller and more homogeneous thickness, smaller mass to volume ratio, and more conic shapes, see Supplementary Section 3.

### Composite arrhythmic outcome analysis

Three PCA modes were discriminative of patients who met the composite arrhythmic outcome: modes 24 (p= 0.006), 14 (p= 0.008) and 25 (p = 0.026), see Figure 3. In a sub-group analysis, mode 24 was the most consistent to differentiate outcomes across groups (p = 0.058 for the iDCM group with 4 events, and p = 0.086 in the aDCM group with 8 events). The LASSO LDA selected these three modes (14, 24 and 25) reaching a high degree of discrimination between the two groups (outcome vs. no outcome, AUC of 0.878 in cross-validation).

**Figure 3:**
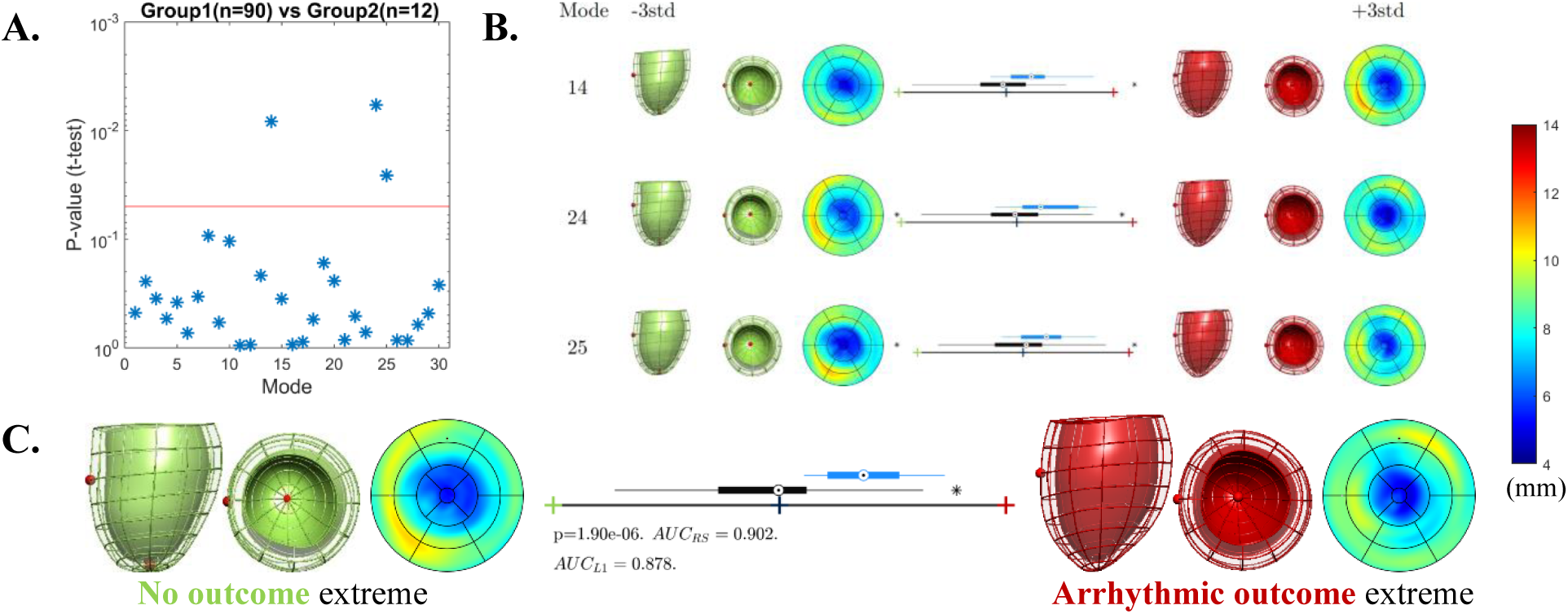
Shape differences captured between patients with (n=12) and without (n=90) arrhythmic composite outcome. **Panel A**: Result of the T-test comparing the two groups, where modes 14, 24 and 25 are above the threshold of significance set at p=0.05. **Panel B**: Illustration of modes 14, 24 and 25 by their extreme shapes (red shape as the extreme associated with the arrhythmic composite outcome and green to the opposite) with corresponding thickness plots and with boxplots of the distribution of the group without outcome (black) and with outcome (blue). **Panel C**: Illustration of the changes encoded by the LDA axis over the first 25 PCA modes, that selected modes 14, 24 and 25 illustrated in Panel B, with boxplots following the same color conventions as in panel B.

Interpretation of the features captured by tthese modes is drawn by the visualization of the extreme shapes in Figure 3. A spiral septal increase in relative wall thickness pattern is the main protective feature that emerged from mode 24 (the AUC of mode 24 was 0.729), as illustrated in panel C of Figure 3. The presence of a conical (i.e. wider basal and mid vs. apical diameter) LV morphology was the main adverse characteristic.

## Discussion

The absence of a spiral septal morphology identified by mode 24 was associated with adverse arrhythmic outcomes and this was a predominant feature of the aDCM subgroup. Besides, a conical LV with thinner walls was characteristic for the aDCM group, as opposed to the iDCM group with relatively thicker walls and a rounder apex shifted towards the RV. These LV remodeling phenotypes suggest novel mechanistic insights and may help in the risk stratification of DCM subjects.

### Robustness of computational modelling and comparison to previous findings

Our methodology explored LV shape beyond traditional bulk metrics such as volume or mass. It uses smooth computational meshes that minimize the impact of potential artefacts such as slice shifts in SAX acquisitions or segmentation noise (16), that reported average fitting error smaller than voxel size (1.18 ± 0.47 mm), and that have been used in a number of previous studies (19,20).

This computational parameterization of cardiac anatomy was similarly employed in 156 patients with exclusively fibrotic DCM to investigate shape in LV remodeling for arrhythmia prediction (12). The adverse prognostic shape derived in that study had a paraboloidal profile (i.e. wider basal to mid cavity diameter) that related to the presence of LGE-related fibrosis. In our study, the extension to a clinically defined, higher risk subgroup of DCM showed a qualitatively similar wider basal and mid to apical cavity diameters (i.e. conicity that differentiated aDCM vs. iDCM, see Figure 2), a shape pattern that appeared independent of the presence of LGE (see Supplementary Fig.2). A more conical morphology is also qualitatively present in the extreme morphology that was linked to a higher risk of arrhythmic outcomes (see Figure 3).

A spherical left ventricle has been described in studies as the main adverse remodeling pattern in iDCM associated with poor survival including heart failure (21). Our results do not reveal any quantitative sphericity metric to be associated to adverse outcomes (see Supplementary Table 3), and as such we interpret the features identified in our study (i.e. conicity, LV axis shift, relative thickness differences in a septal spiral morphology) are linked to those of an earlier clinical phenotype or a unique electrophysiological remodeling pattern.

### Phenotypic heterogeneity in DCM

The iDCM subgroup had greater volumes, ventricular mass, and a more advanced disease as evidenced by the lower EF (median LVEF 34% in iDCM vs. 40% in aDCM subgroups). These features are acknowledged as part of the known spectrum of adverse remodeling in DCM, associated with redistribution of wall stress and fiber shortening affecting resting contractile function over time (21–23).

The aDCM subgroup represent a subset of familial DCM that is distinct from other forms of DCM (2). This is appreciated in this study by the higher average LVEF and higher baseline arrhythmic profile. Up to 22% of aDCM patients had LV regional WMA, highlighting the possibility of specific arrhythmic genotypes such as the appreciated Desmoplakin (DSP) in this study and Filamin C (FLNC), both of which have been described within the current concept of left dominant ACM (2,24). Whilst regional WMA may be present in aDCM patients, they are not specific enough to identifying this subgroup (capturing less than a quarter of the population in this study), nor do they directly relate to underlying arrhythmic risk which can predate onset of WMA (24). Supplementary section 4 reports how WMA was not a significant confounder factor on the LDA score that differentiated these subgroups.

### Potential arrhythmogenic mechanisms driven by LV shape

Presence of thicker walls in a spiral septal morphology (i.e. mode 24) was the novel shape feature identified that was associated with iDCM and with a decreased arrhythmic risk, in both cases with the strongest significance across all modes (see Figure 2 and Figure 3). This feature discriminated the risk across the two sub-groups (aDCM and iDCM) with similar performance. This raises the mechanistic question of why this pattern would be more protective against arrhythmic risk.

This study did not provide evidence that this localized septal wall morphology related to the process of focal reparative fibrosis. Mode 24 did not discriminate the presence or not of LGE in our cohort. Moreover, presence of LGE was not a confounder for the LDA risk score (see Supplementary Section 4). This, however, does not exclude the contribution from more concealed areas of diffuse, interstitial fibrosis, also recognized in the histopathology of DCM, that will be overlooked by use of LGE-imaging alone (25,26). Thus, it is still plausible that the lack of this septal spiral morphology could signal areas of higher wall stress from diffuse fibrotic processes. Further studies incorporating T1 mapping, a non-invasive index of diffuse fibrosis, and electrophysiological parametric mapping could investigate this hypothesis.

Another plausible mechanistic link is that the spiral pattern signals an adaptive remodeling to increased workload, and as such it signals the presence of healthy cell behavior (i.e. compensatory myocyte thickening). On the contrary, those cases that do not respond (do not thicken as cavity dilates) show a lack of adaptive ability – this was the mechanistic link to explain the ability to predict responders to CRT by an asymmetric wall thickening pattern (19). In our case, the spiral pattern identified starts at the outflow tract, and travels through the septum, the flattest anatomical part of the left ventricle and thus holding larger stress by Laplace’s law (the flatter the surface, the larger the stress) (27).

A second important consideration is that the spiral pattern found suggests a plausible link to the role of helical fibers in the development of effective ejection fraction (28): it suggests the presence of aligned cardiomyocyte orientations undergoing a high workload. Related to this, geometric abnormalities that cause a loss of cardiomyocyte re-orientation during contraction have been reported to underlie the decrease of LVEF in eccentric hypertrophy patterns of DCM (29). Our data agrees with this observation, since mode 1 captured the progression to eccentric hypertrophy (gradual increase of volume with linked wall thickening and increased sphericity) displayed a very strong negative correlation with LVEF (coefficient of determination R^2^ of a 45.6%). We thus hypothesize that in DCM there is a loss of cardiomyocyte orientation (e.g. that may be caused by the presence of diffuse fibrosis) that causes the loss of the ability to develop further work (extra workload that would manifest with a mild spiral septal hypertrophy), and that this population is associated with a larger arrhythmogenic risk.

Another important consideration is the presence of spiral septal morphology in some genetic forms of hypertrophic cardiomyopathy (HCM), that are qualitatively similar to those of mode 24 but of much larger amplitude (20). The question is then why HCM develops such extreme phenotype while DCM would develop a much milder version. In HCM the impaired contraction has been linked to the loss of cardiomyocyte re-orientation during relaxation, and not during contraction as in DCM (29) . We then refine the previous hypothesis that the impaired cardiomyocyte re-orientation during contraction, and not during relaxation, is the one associated with the loss of the spiral septal morphology and with a larger arrhythmic risk.

It was also not possible to fully exclude genetic contributions. FLNC or DSP pathogenic variants have characteristically more regionality in LV impairment, which was prominent in approximately 20% of our aDCM cohort (24). From a multicenter experience of 25 consecutive Lamin A (LMNA)-mutation patients undergoing ablation of sustained VT, the VT predominantly related to re-entry in and around regions of scar located mostly in the basal septum and is described as spiraling towards an LV exit (30). In a separate study of 15 patients with a Titin (TTN) cardiomyopathy, intramural circuits that embedded extensively within the septal and perivalvular regions formed the most common substrate distribution for those with ventricular arrhythmias (31).

### Clinical perspectives: risk stratification in DCM

Genotyping and fibrosis assessment have clear benefits and show promise in risk stratification in DCM (32). However, due to phenotypic heterogeneity and lack of clear genotype-phenotype correlations for multiple variants, accurate characterization of arrhythmic risk in DCM is still a major challenge. In this study, we have discovered a morphological phenotype, a mild spiral septal thickening pattern, that differentiated arrhythmic risk with consistent performance across the two subgroups (iDCM and aDCM), and that is interpreted to be a common protective feature across DCM that could improve risk stratification. Furthermore, this remodeling pattern (LDA score) remained robust to demographic factors including age, which is widely recognized as a potential confounding factor for assessing adverse cardiovascular outcomes (See Supplementary Section 4).

### Study Limitations

The observational nature of the study analysis offers exploratory associations to be made between novel shape metrics and an arrhythmic DCM population. To minimize the impact from prior referral bias, individuals were consecutively recruited according to expert consensus and CMR assessment. An external cohort is needed for validation of these initial findings given the relatively small sample size. Furthermore, the shape modes that were discriminative between ADCM and IDCM and the event vs. no-event groups explained relatively small fractions of the total shape variance, hence they represented subtle shape features which should be further investigated in external cohorts.

Secondly, the aDCM subgroup were selected on clinical consensus grounds of future increased arrhythmic risk, however the study was only evaluated after a 5-year follow up period and therefore the arrhythmic event rate during this time was limited. This potentially conflicts the weight of the number of arrhythmic endpoint outcomes achieved and thereby the differences between subgroups.

Finally, whilst every effort was made to exclude competing comorbidities that could also contribute to a DCM phenotype, it is plausible that some patients in the iDCM subgroup had overlapping hypertension as an aggravating factor. This could contribute to the suggested and relative hypertrophy pattern seen in this subgroup. Reassuringly, there were no features of hypertensive heart disease identified as part of these routine clinical CMR studies, but a larger validation study with comparison to an exclusive cohort of those with this phenotype would better clarify this position.

## Conclusions

The lack of a spiral septal morphology was associated with both the familial subset (aDCM) and an increased arrhythmic risk. This may serve as an additive predictor for risk stratification in this subpopulation with a propensity towards malignant arrhythmias and poor prognosis that is not entirely explained by LV dysfunction alone. Further research is warranted to clarify this position and better understand the electrophysiological basis for these findings and the relationship to genetic substrates.

## Ethics statement & Consent

All participants provided written informed consent. This study protocol was approved by the North of Scotland Research Ethics service (REC approval number: 15/NS/0030) and conducted in accordance with the Declaration of Helsinki.

